# Polyester Nasal Swabs Collected in a Dry Tube are a Robust and Inexpensive, Minimal Self-Collection Kit for SARS-CoV-2 Testing

**DOI:** 10.1101/2020.10.09.20210302

**Authors:** Leah R. Padgett, Lauren A. Kennington, Charlotte L. Ahls, Delini K. Samarasinghe, Yuan-Po Tu, Michelle L. Wallander, James S. Elliott, Douglas Rains

**Affiliations:** Quantigen Biosciences, Fishers, IN, United States; The Everett Clinic-Part of Optum, Everett, WA, United States; Sciest LLC, Mooresville, NC, United States

**Keywords:** SARS-CoV-2, nasal swabs, dry swabs, polyester swabs, stability, RT-qPCR

## Abstract

**Background:** Polyester nasal swabs stored in saline or in a dry tube were evaluated as an alternative to foam nasal swabs for SARS-CoV-2 testing by reverse transcription quantitative polymerase chain reaction (RT-qPCR) since they may be inexpensively manufactured at high capacity.

**Methods:** Surrogate clinical specimens were prepared by inoculating foam and polyester nasal swabs with residual SARS-CoV-2 positive clinical specimens diluted in porcine or human matrix. Dry swab elution with phosphate buffered saline (PBS) was evaluated by vortex, swab swirling, and passive methodologies. Surrogate and clinical nasal specimen stability were evaluated at refrigerated (4°C) and elevated temperatures (40°C for 12 hours, 32°C hold) through 72 hours.

**Results:** Polyester swabs demonstrated equivalent performance to foam swabs for detection of low and high SARS-CoV-2 viral loads. Dry swab elution performed with PBS and mechanical disruption by vortex resulted in nearly complete quantitative recovery of virus. Dry polyester and foam surrogate specimens were stable through 72 hours both when refrigerated and after high temperature excursion, which simulated specimen transport without cold chain. Similarly, clinical specimens collected with polyester swabs and stored dry were stable through 72 hours in the presence and absence of cold chain. Polyester surrogate specimens stored in saline were stable through 72 hours refrigerated but only through 48 hours at elevated temperatures.

**Conclusions:** Polyester nasal swabs stored in dry collection tubes comprise a robust and inexpensive self-collection method for SARS-CoV-2 viral load testing, which is stable under conditions required for home collection and shipment to the laboratory.

## Introduction

The worldwide outbreak of severe acute respiratory syndrome coronavirus-2 (SARS-CoV-2) has caused unprecedented demand for testing to diagnose patients with coronavirus disease 2019 (COVID-19). To expand capacity and increase accessibility, nasal (anterior nares, AN) specimens were evaluated for SARS-CoV-2 molecular testing and were deemed acceptable by the FDA, leading to inclusion in recommendations by both the CDC and the FDA (1, 2). Recent studies demonstrated that self-collected AN specimens are equivalent to healthcare-professional collected nasopharyngeal and oropharyngeal swabs for detection of SARS-CoV-2 by reverse transcriptase-polymerase chain reaction (RT-qPCR) (2, 3). Nasal swab self-collection is more tolerable and reduces patient contact with medical professionals, thereby decreasing the risk of viral transmission and preserving PPE supplies.

In early March of 2020, the initial studies which established the nasal swab collection method utilized foam swabs (2, 3). However, supplies quickly became depleted as large diagnostic manufacturers, laboratories, and government agencies secured the available supply. To address the shortage, additional swab types were evaluated including spun polyester swabs, which can be manufactured at higher capacity than foam swabs and are inexpensive to produce. The polyester spun swab demonstrated similar performance for nasal specimen collection to the foam swab and FDA guidance was updated to include polyester swabs as an acceptable swab type for SARS-CoV-2 testing (1, 4).

Upper respiratory swabs have historically been transferred to the laboratory in viral transport media (VTM), however VTM shortages ensued upon the start of the pandemic. Phosphate buffered saline (PBS), saline, and other alternative solutions were deemed acceptable by the FDA for swab transport (1), with saline demonstrating better performance for use with nasal swabs as compared to VTM (4). While alternate swab transport solutions have alleviated the immediate supply shortages, consideration of additional swab transport methods such as dry tube storage prompted further investigation.

In this study, we evaluated the performance and stability of polyester nasal swabs stored in dry collection tubes as an alternative specimen collection option for SARS-CoV-2 molecular testing. Replicate swabs were prepared using residual positive clinical specimens diluted in clinical matrix to known concentrations near the limit of detection (LoD). The conclusions of these studies were confirmed by the assessment of fresh positive patient specimens collected with polyester nasal swabs. We demonstrate that clinical specimens stored dry are stable under conditions required for home collection and shipment to the laboratory for testing.

## Materials and Methods

### SARS-CoV-2 Clinical Specimens

Residual clinician-collected nasal specimens were collected in sterile saline or VTM and refrigerated (UnitedHealth Group protocol #2020-0002, approved by UHG Institutional Review Board). All positive specimens were collected from known COVID-19 patients previously tested using the TaqPath COVID-19 Combo Kit (Thermo Fisher Scientific) according to the Emergency Use Authorization (EUA) instructions (5).

### Source Materials

Nasal swabs included: Puritan foam (#25-1506), Fisherbrand polyester-tipped applicator (Fisher Scientific, #22-363-170), Copan spun polyester (#164KS01), SteriPack spun polyester #60564 (also known as US Cotton spun polyester #3), SteriPack spun polyester #60567 (also known as US Cotton spun polyester 3ARS), Copan mini FLOQ (#503CS01), and Copan regular FLOQ (#56380CS01). PBS (Sigma) was used for dry swab elution and sterile saline (Kroger) was used for swab storage when media was utilized. Porcine mucus (clinical matrix) was obtained from the University of Minnesota Veterinary Diagnostic Lab and the Iowa State University Veterinary Medicine Laboratory.

### Dry Swab Elution

Dry swab elution was performed by adding 1 mL of PBS to the 15 mL screw top tube, a 30 second vortex with intermittent pulsing, and incubation for 10 minutes at room temperature prior to RNA extraction.

### RNA Extraction and RT-qPCR

RNA extraction was performed from 400 µL PBS eluate using the MagMAX™ Viral/Pathogen Nucleic Acid Isolation Kit (Thermo Fisher). RT-qPCR was performed on the QuantStudio™ 7 Flex 96-well Fast (Thermo Fisher) using 5 µL RNA and the TaqPath COVID-19 Combo Kit according to the EUA Instructions For Use, 14March2020; Pub. No. MAN0019181 Rev. A.0 (5). Raw cycle thresholds were obtained using the QuantStudio™ Real-Time PCR Software v1.3 and the following thresholds: 40,000 for *ORF1ab*, 25,000 for *N*, and 38,000 for *S*. Samples with indeterminate or spurious amplification signals were designated a cycle threshold (Ct) value of 40 cycles. For the experiments using human matrix, RNA samples were also tested using the TaqMan™ RNase P assay (Fisher Scientific) on the QuantStudio™ Dx Real-Time Instrument.

### Construction of Surrogate Clinical Specimens

Residual specimens with negative SARS-CoV-2 results were pooled and diluted 1:2 in porcine mucus to generate the negative clinical matrix. Twelve high-positive SARS-CoV-2 specimens with Ct values < 25 cycles for all three viral target genes (*ORF1ab, N, S*) were combined to generate a master positive pool and replicates were tested to determine the average Ct values. Ct values corresponding to 2x LoD (20 genomic copy equivalents (GCE)/PCR reaction) and 10x LoD (100 GCE/PCR reaction) were calculated using the formula 2^-ΔCt^=F, where ΔCt is the necessary Ct adjustment and F is the fold change (6). Using *N* gene ΔCt calculations, the master positive pool was diluted with negative clinical matrix to generate two positive clinical pools; 2x LoD (low-positive pool) and 10x LoD (high-positive pool). Ten replicates of the low-and high-positive clinical pools were tested to confirm that *N* gene Ct values were within the expected range (± 20% linear fold change). SARS-CoV-2 GCE values for the clinical pools were determined by *N* gene standard curve analysis and digital PCR (**Supplemental Data**).

### Viral Recovery from Swabs

Viral recovery from polyester swabs (Copan) was evaluated across a range of viral loads (1x to 64x LoD) by serial dilution of the master positive specimen pool with negative specimen clinical matrix. The viral pools were tested directly (no swab) or spiked onto polyester swabs. To create the spiked swabs, polyester swabs were submerged into a 1.5 mL tube containing 40 µL of positive pool and abraded against the tube wall five times in each direction to mimic nasal collection or until all of the specimen material was absorbed. The inoculated swab was then transferred to a 15 mL screw cap tube. For the direct viral testing, 40 µL of positive pool was aliquoted into a 15 mL screw cap tube. Three replicates were performed for each of the clinical positive pools and conditions (+/-swab). All tubes, with or without swabs, were incubated for 30 minutes at room temperature followed by dry swab elution with 1 mL PBS for the spiked swab or addition of 1 mL PBS directly to the tube (no swab).

For the evaluation of alternative swab elution methods, paired polyester swabs (SteriPack #60567) were collected from the AN of 10 healthy volunteers by an equal loading method with a crossover design (7). Negative specimens were spiked with 40 µL of the low-positive (2x LoD) clinical specimen pool and stored at 4°C in a dry tube overnight. Swabs were divided into two groups, each containing five pairs, and eluted by the addition of 1 mL PBS. Group 1 compared the standard vortex method with swab swirling (swirling the swab within the tube for 10 seconds) while group 2 compared the standard vortex method to passive elution for 1 hour under refrigeration. RNA was extracted for SARS-CoV-2 and RNase P testing by RT-PCR.

### Evaluation of Surrogate Specimen Stability (Porcine Matrix)

A two-arm study (**Supplemental Figure 1**) was performed to evaluate the stability of surrogate clinical specimens prepared using foam (Puritan) and polyester swabs (Fisherbrand) stored in dry tubes, as well as polyester swabs (Fisherbrand) stored in a tube containing 1 mL of sterile saline. Swabs were stored at 4°C (control arm) or at 40°C for the initial 12 hours and then held at 32°C (experimental arm). To create the surrogate clinical specimens, 2x LoD and 10x LoD specimen pools were added directly to the swabs through a procedure that mimicked nasal swabbing action. Each swab was placed in a 1.5 mL microcentrifuge tube containing 80 µL of the appropriate specimen pool, with the tip of each submerged into the mixture and abraded in a circular motion against the side of the tube for 30 seconds. Swabs were transferred to 15 mL screw cap tubes containing either 1 mL saline or with no addition of media (dry). After inoculation, baseline and negative (n=2 per swab type) swabs from the control arm were processed immediately (0 hours). Remaining swabs were stored at 4°C or 40°C for the control and experimental arms, respectively. After 12 hours at 40°C, experimental swabs were transferred to 32°C. Swab stability was assessed at three time points: 1) 24 hours (+ 8 hours), 2) 48 hours (+ 8 hours), and 3) 72 hours (+ 8 hours). The additional eight hours were included as a FDA requirement for stability labeling. Swabs were retrieved from storage two hours prior to each time point to equilibrate to room temperature.

**Figure 1.**
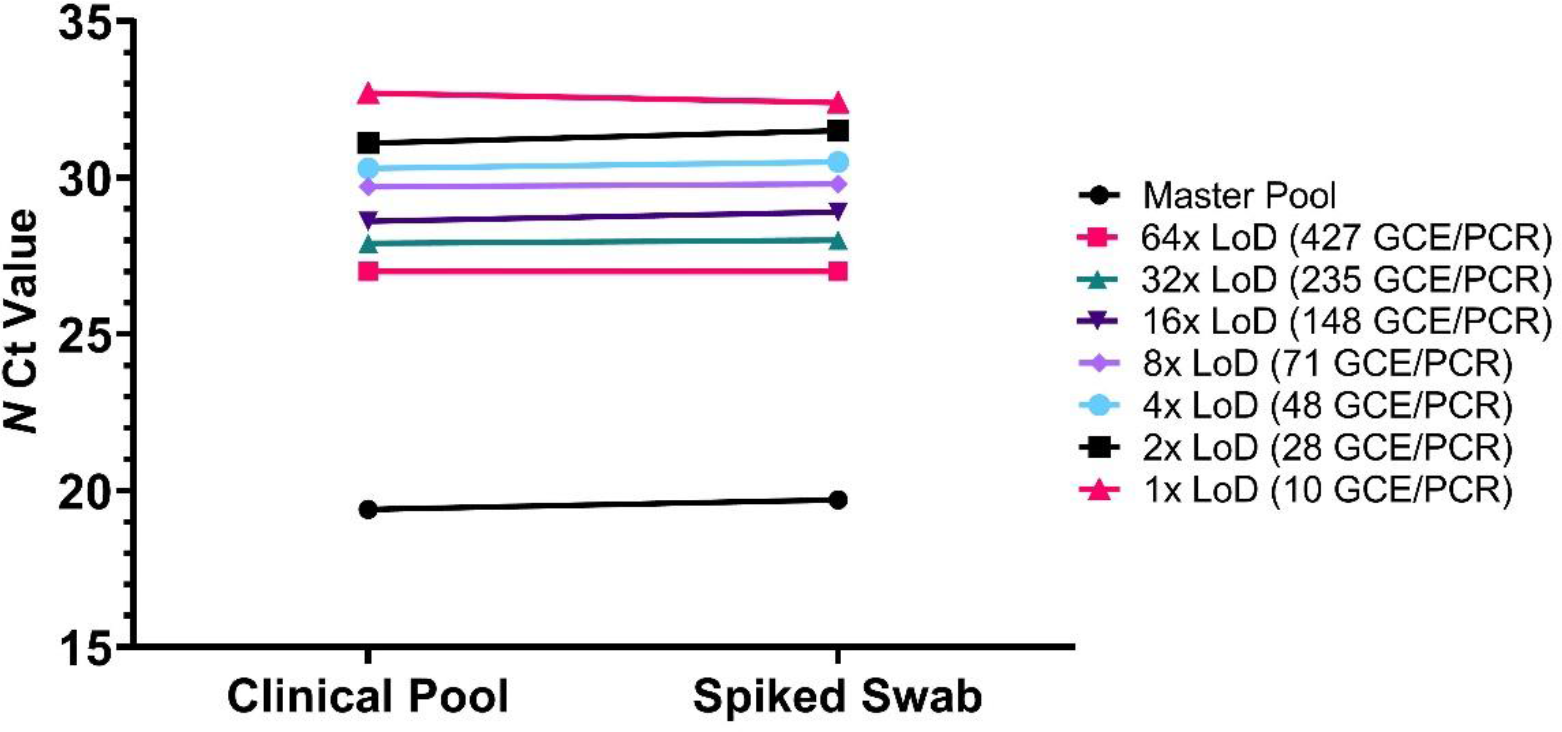
Evaluation of absorption and recovery from dry polyester swabs across a range of viral loads. *N* gene mean Ct values from clinical pool serial dilutions (n=3) tested directly were compared to *N* gene mean Ct values from swabs spiked with clinical pool serial dilutions (n = 3).

### Evaluation of Polyester Swab Clinical Specimen Stability

Dry swab specimen stability using human matrix was evaluated through 72 hours at 4°C and through 48 hours at elevated temperatures (40°C 12 hours, held at 32°C). Healthy volunteers were each given one Copan polyester swab and one SteriPack #60564 polyester swab and were instructed to collect normal AN clinical matrix onto the swabs to generate roughly equivalent replicates as follows: 1) five revolutions of the Copan swab (left nostril), 2) five revolutions of the SteriPack #60564 swab (right nostril), 3) five revolutions of the same SteriPack #60564 swab (left nostril), and 4) five revolutions of the same Copan swab (right nostril) (7). The swabs were collected from volunteers within an eight-hour timeframe and stored overnight in 15 mL screw cap tubes at 4°C prior to viral inoculation. Each swab was placed in a 1.5 mL microcentrifuge tube containing 40 µL of one SARS-CoV-2 high-positive (10x LoD) patient sample, with the tip submerged into the mixture and abraded in a circular motion against the side of the tube for 30 seconds or until complete absorption. Swabs were transferred to dry 15 mL screw cap tubes. Baseline swabs were processed and tested immediately (0 hours), while all remaining swabs were stored at 4°C or 40°C for the control and experimental arms, respectively. After 12 hours at 40°C, experimental swabs were transferred to 32°C. Specimens were retrieved from storage two hours prior to each time point and were allowed to equilibrate to room temperature. Specimen stability was assessed at 24 hours (+ 8 hours) and 48 hours (+ 8 hours) in both study arms and at 72 hours (+ 8 hours) in the control arm.

For the evaluation of clinical specimens, paired SteriPack polyester #60564 swabs were collected from five SARS-CoV-2 positive and five SARS-CoV-2 negative patients by the same method described above. Paired dry swabs were stored at 4°C or 40°C for the control and experimental arms, respectively. After 12 hours at 40°C, experimental swabs were transferred to 32°C. Specimens were retrieved from storage two hours prior to each time point and were allowed to equilibrate to room temperature. Specimen stability was assessed at 72 hours (+ 8 hours).

### Data Analysis

Specimen stability at each time point was defined as ≥ 95% agreement with the expected result for low-positive samples (2x LoD) and ≥ 100% agreement with the expected results for high-positive (10x LoD) and negative control samples. When calculating the differences between Ct values, negative ΔCt values indicate a lower Ct and thus higher viral load, while positive ΔCt values indicate a higher Ct value and thus lower viral load. All statistical analyses (one-way ANOVA and t-test) were performed using GraphPad Prism 8.

## Results

### Swab Absorption and Elution from Swabs Stored in Dry Format

Surrogate specimens were prepared by spiking polyester and foam nasal swabs with SARS-CoV-2 positive clinical specimen pools using a swab swirling procedure to mimic nasal swabbing. Polyester swabs were 150% more absorbent than foam swabs by the swirling method and by passive absorption (**Table 1**). Once prepared, surrogate specimens were stored dry in empty tubes and eluted with 1 mL PBS, followed by a 30 second vortex and 10 minute room temperature incubation. Specimen recovery from dry swabs was evaluated by comparing Ct values from serially diluted surrogate specimens created with polyester swabs to Ct values from serially diluted clinical specimen pools tested directly, without absorption and elution from swabs. SARS-CoV-2 *N* gene mean Ct values from swabs and pools were not statistically different from one another (p>0.05) for viral loads ranging from 10 to 66,779 GCE/PCR reaction (**Figure 1**). Mean ΔCt values were < 0.5 cycle indicating that there is nearly complete quantitative absorption and recovery of clinical specimens from swabs using the vortex elution method.

**Table 1.** Swab dimensions and average absorption volume (n = 10).

| Swab | Average<br>Length (mm) | Average<br>Width (mm) | Average Absorption (μL) |  |
| --- | --- | --- | --- | --- |
|  |  |  | Passive<br>10 sec | 5 swirls in<br>Each Direction |
| Puritan Foam | 15.4 | 4.5 | 28 | 85 |
| Fisherbrand Polyester | 13.7 | 5.0 | 121 | 112 |
| Copan Polyester | 15.3 | 4.8 | 113 | 120 |
| SteriPack Polyester (#60564) | 16.1 | 5.2 | 182 | 184 |
| SteriPack Polyester (#60567) | 15.1 | 4.5 | 120 | 150 |
| Copan FLOQ | 25.5 | 3.8 | 24 | 64 |
| Copan Mini-FLOQ | 15.5 | 3.8 | 50 | 23 |

Two additional dry swab elution methodologies, swab swirling within the tube and passive recovery, were compared to the vortex method to determine if mechanical disruption is necessary for full specimen recovery. Paired polyester swabs collected from healthy volunteers were spiked with a low-positive clinical specimen pool (2x LoD) and dry swabs were eluted using the vortex and swab swirling methods or the vortex and passive methods. Mean Ct values for the SARS-CoV-2 targets (*N, ORF1ab*, S) and human *RNase P* were all statistically higher (p<0.05) for swirled and passively eluted swabs than the paired vortexed swabs (**Figure 2**). Mean paired ΔCt values ranged from one to three cycles, correlating to a two-to seven-fold reduction in sample detection. These results indicate that mechanical disruption by vortex is necessary for full elution of viral material from dry swabs.

**Figure 2.**
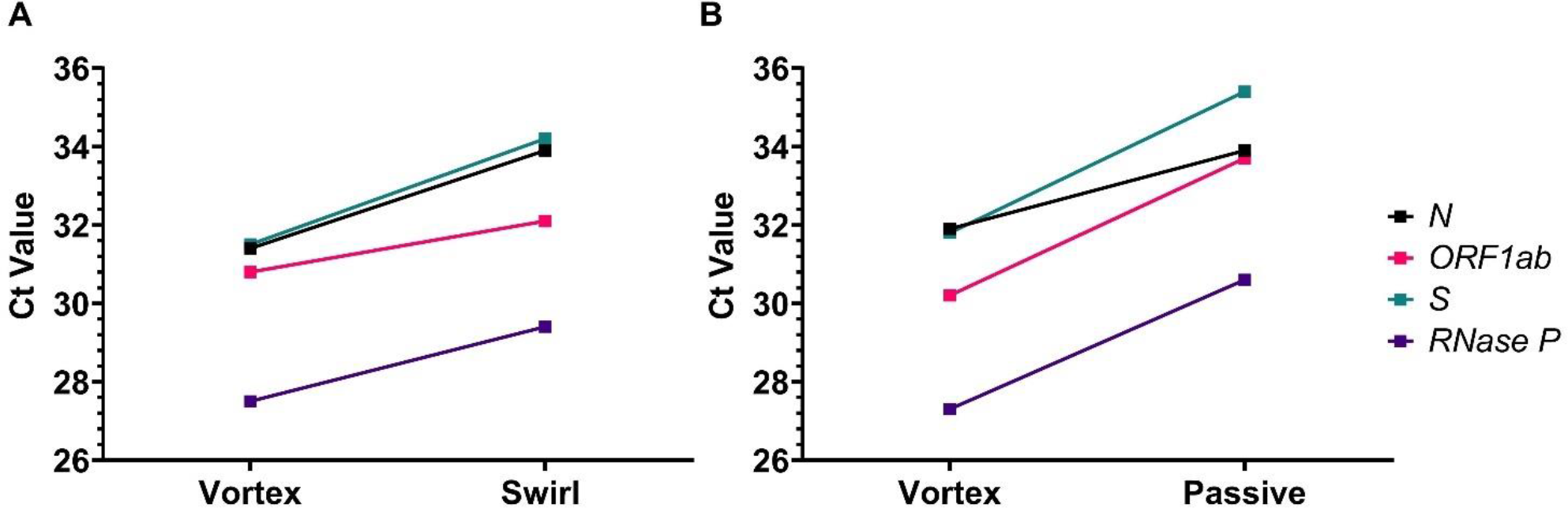
Comparison of the dry swab vortex elution method with swab swirling (**A**) and passive elution (**B**). Mean Ct values are shown from paired polyester swabs collected from human volunteers (n=5) and spiked with the low-positive clinical pool (2x LoD; 20 GCE/PCR reaction).

### Polyester and Foam Swab Surrogate Specimen Stability

Stability of swabs for RT-qPCR detection of SARS-CoV-2 was assessed with and without cold chain storage. Polyester and foam swabs stored dry (“dry polyester” and “dry foam”) and polyester swabs stored in saline (“polyester saline”) were prepared using low-or high-positive SARS-CoV-2 clinical specimen pools containing 22 and 144 GCE/PCR reaction, respectively. Ct values from the three swab types held under refrigerated conditions were not statistically different (p>0.05) through 72 hours (**Figure 3A**). Refrigerated dry polyester, dry foam, and polyester saline low-and high-positive specimen replicates had 100% positive agreement, demonstrating stability through 72 hours. At elevated temperatures (40°C 12 hours, 32°C hold), all three swab types had 100% positive percent agreement for high-positive specimen replicates through 72 hours. Ct values for low-positive dry polyester and dry foam swab specimens were not statistically different (p>0.05) through 72 hours, indicating that the two swab types were equivalent (**Figure 3B**). For both, one of 20 low-positive specimen replicates were inconclusive (only *N* detected) after 72 hours at elevated temperatures, resulting in 95% positive percent agreement, meeting the study acceptance criteria and establishing stability for dry specimens through 72 hours. Stability for polyester saline specimens was established through 48 hours at elevated temperatures (100% positive agreement), however, Ct values were significantly higher (p<0.05) than dry polyester and foam after 48 hours. Seven of 20 low-positive polyester saline replicates were inconclusive (only *N* detected) after 72 hours at elevated temperatures for 65% positive percent agreement; thus, stability for polyester saline specimens was established through 48 hours at elevated temperatures (100% positive agreement).

**Figure 3.**
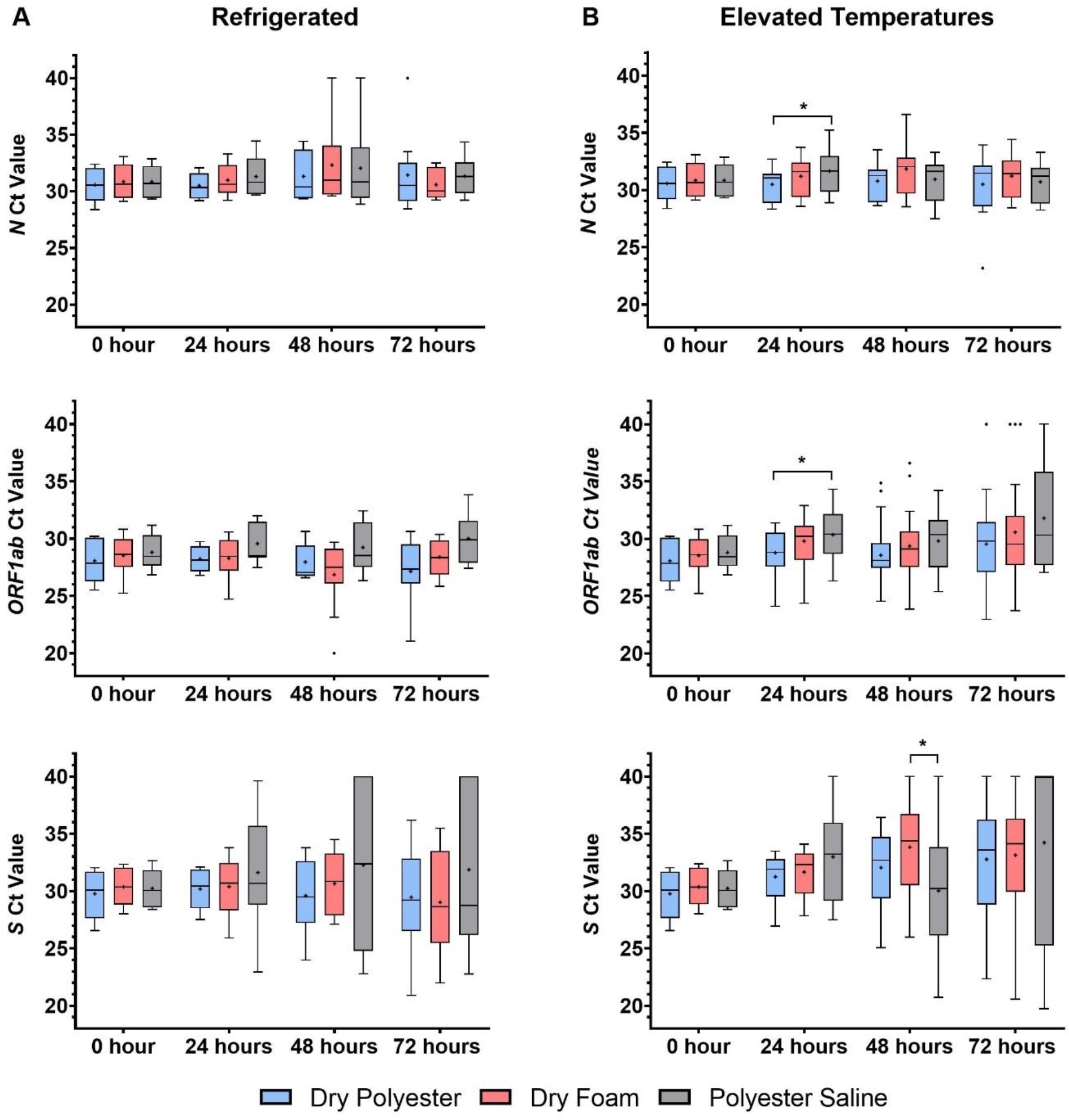
Quantitative measurement of nasal swab specimen stability. Dry polyester (blue), dry foam (red), and polyester swabs stored in saline (gray) were stored refrigerated (**A**, 4°C) or at elevated temperatures (**B**, 40°C 12 hours, 32°C) through 72 hours. No PCR amplification was assigned a Ct value of 40. *statistically different, p<0.05.

Viral RNA levels were relatively stable for the dry swabs through 72 hours at both temperatures, with ΔCt values for *N* and *ORF1ab* < 3 cycles (less than 10-fold change in viral RNA levels) (**Table 2**). Polyester swabs in saline were also stable through 72 hours when refrigerated, however they were less stable at elevated temperatures and only met the acceptance criteria through 48 hours. The *S* gene target demonstrated diminished stability for all three specimen types at elevated temperatures and extended time periods when refrigerated, with less relative stability in saline versus dry. The detection of only two out of three viral targets were required for a positive SARS-CoV-2 result in accordance with the interpretation algorithm, therefore the *S* gene results did not impact the stability claims for the dry polyester and foam swabs. The Ct drift analysis confirmed that at elevated temperatures, dry polyester and dry foam specimens are stable through 72 hours with ΔCt values < 3 cycles while polyester specimens stored in saline are stable through 48 hours.

**Table 2.** Changes in viral load during time and temperature stability studies. Mean ΔCt values (stability time point – 0 h) for low-positive (2x LoD) samples. Specimens with indeterminate results or no amplification were assigned 40 Ct cycles. Bold indicates ΔCt > 3.0 cycles.

| SARS-CoV-2 Target | Swab | Refrigerated ΔCt | | | Elevated Temperatures ΔCt | | |
| --- | --- | --- | --- | --- | --- | --- | --- |
|  |  | 24 h | 48 h | 72 h | 24 h | 48 h | 72 h |
| *N* | Dry Polyester | -0.5 | 1.1 | 1.8 | -0.7 | -0.6 | -0.6 |
|  | Dry Foam | -0.1 | 2.5 | -0.8 | -0.2 | 0.7 | -0.1 |
|  | Polyester Saline | 0.6 | 2.5 | 0.3 | 0.5 | -0.2 | -0.7 |
| *ORF1ab* | Dry Polyester | -0.4 | -0.5 | -1.3 | -0.1 | -0.6 | 1.4 |
|  | Dry Foam | -0.1 | -2.0 | -0.1 | 1.0 | 0.2 | 2.0 |
|  | Polyester Saline | 0.9 | 1.0 | 1.9 | 1.3 | 1.1 | **3.9** |
| *S* | Dry Polyester | 0.2 | 0.7 | 1.4 | 0.7 | 1.3 | **3.9** |
|  | Dry Foam | 0.6 | 1.4 | 1.0 | 1.0 | **4.1** | **4.2** |
|  | Polyester Saline | **3.7** | **7.6** | **6.2** | **3.3** | 0.7 | **7.4** |

### Polyester Swab Human Matrix and Clinical Specimen Stability

An additional evaluation of dry polyester swab specimen stability was performed using matrix from individual human volunteers. Paired Copan and SteriPack #60564 polyester nasal swabs were self-collected by healthy adults and inoculated with virus to create high-positive (10x LoD) specimens. Consistent with the results obtained with surrogate specimens prepared with porcine matrix, in the presence of human matrix, contrived dry polyester swabs were stable for SARS-CoV-2 detection through 72 hours refrigerated and through at least 48 hours at elevated temperatures (the experiment was only conducted to 48 hours + 8 hours) (**Supplemental Table 1**).

The stability of contrived dry swab specimens was confirmed with clinical specimens collected from five SARS-CoV-2 positive individuals and five healthy individuals using paired polyester swabs. Paired swabs were stored for 72 hours with one swab refrigerated and the other at elevated temperatures. Ct values for the three SARS-CoV-2 targets and *RNase P* were not statistically different (p>0.05) by storage temperature (**Figure 4**), indicating that dry clinical specimens are stable through 72 hours at both refrigerated and elevated temperatures.

**Figure 4.**
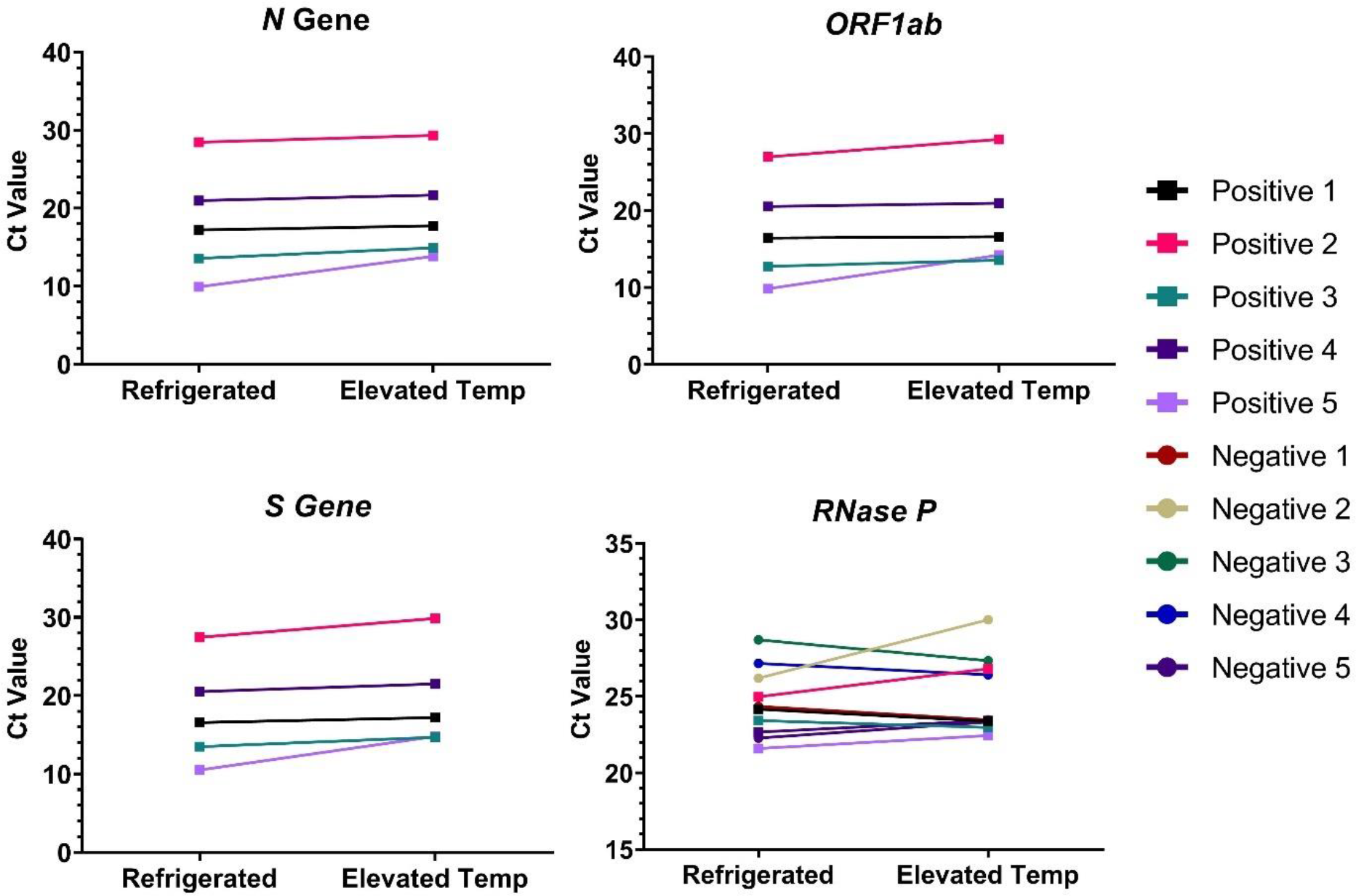
Dry clinical specimen stability. Paired collections from SARS-CoV-2 positive (n=5) and negative (n=5) patients were stored refrigerated and at elevated temperatures though 72 hours prior to RT-qPCR analysis for SARS-CoV-2 and *RNase P*.

## Discussion

Self-collected nasal specimens from the AN perform equivalently to nasopharyngeal and oropharyngeal specimens for SARS-CoV-2 detection by RT-qPCR (2, 3). In this study, we demonstrate that very low viral loads near the assay limit of detection are detectable from nasal swab surrogate clinical specimens that are consistent with viral loads through 10 days from the start of COVID-19 symptoms (*Y-P Tu, personal communication*, August 2020). Viral infectivity studies conducted by the CDC and other groups have shown that specimens collected from confirmed SARS-CoV-2 patients with mild to moderate illness who are not severely immunocompromised and who are greater than 9 – 11 days from the start of symptoms do not yield culturable virus, thereby supporting the notion that nasal swab specimens are sensitive enough for detection of the virus through the infectious phase (8-11). In the present study, we demonstrate the performance of AN swabs for SARS-CoV-2 testing using surrogate and clinical specimens and show that self-collected nasal swab specimens produce sufficient material for molecular testing as demonstrated by the reproducible detection of the *RNase P* human target. Importantly, the self-collection instructions for nasal swabbing must be accurately followed to ensure a quality specimen for molecular testing (12). In a recent usability study for the HealthPulse@home Nasal Specimen Collection Kit, AN specimens were successfully self-collected (n=33) as demonstrated by *RNase P* detection (mean Ct = 26.4) using swabs transported dry in the absence of cold chain storage (*Audere, personal communication*, August 2020). The advantages of self-collected swabs from the AN, whether collected at home or in a healthcare setting, include improved patient tolerance, reduction of healthcare worker risk of infection, and preservation of PPE. It is anticipated that compliance with COVID-19 testing recommendations will increase with the use of a well-tolerated and simple collection device and will enable serial testing when appropriate.

In the present study, we demonstrated that polyester swabs from multiple manufacturers are a suitable alternative to foam swabs for molecular detection of SARS-CoV-2. We evaluated the performance of polyester and foam nasal swabs stored in the absence of any transport media within a dry tube and developed a dry swab elution method utilizing PBS and a brief vortex, which can be easily performed in the laboratory. Mechanical disruption by vortex resulted in nearly quantitative recovery of virus from dry swabs even at very low virus levels, while passive elution and swab swirling showed a marked reduction in viral recovery (although this may be sufficient in certain settings). We show that foam and polyester nasal swab specimens may be stored dry without transport media for up to 72 hours in the absence of cold chain. Polyester swabs stored in saline in the absence of cold chain were stable through 48 hours, however there was reduced stability of the low-positive (2x LoD) SARS-CoV-2 samples at 72 hours. Note that buffered saline, such as PBS, may support extended stability beyond 48 hours.

The use of pre-positioned nasal swab collection kits in the home or workplace could facilitate more rapid COVID-19 testing and reduction in transmission. Differential diagnosis of other upper respiratory infections including influenza, which is also detectable in nasal specimens (13), would be feasible from a self-collected specimen. The nasal swab specimens can be stably transported in dry tubes within 72 hours to the laboratory where they can be eluted using a robust and reproducible elution method. Studies to evaluate reduced elution volumes for increased assay sensitivity or direct swab elution into extraction or PCR buffer are under way. The utility of dry swabs can also be extended to future applications, including direct antigen testing for SARS-CoV-2. It is hoped that the inexpensive polyester nasal swab will bolster testing capabilities to improve patient outcomes specifically in low resource settings where swabs and other testing components are in limited supply.

## Data Availability

Upon request, the authors will make any and all data from the paper freely available to members of the research community.

## Acknowledgments

Many thanks to Karen Heichman (Bill & Melinda Gates Foundation) for discussions and perspectives on study design. We thank Bob Setterquist (ThermoFisher), Thomas Bakken (University of Minnesota), Laura Bradner and Karen Harmon (Iowa State University), and Shawna Cooper and Paul Isabelli (Audere) for providing materials and reagents. We thank the Everett Clinic-Part of Optum and UnitedHealth Group for clinical specimens.

## List of Abbreviations

AN: anterior nares
RT-qPCR: reverse transcription quantitative PCR
PBS: phosphate buffered saline
CDC: Centers for Disease Control and Prevention
FDA: U.S. Food and Drug Administration
PPE: personal protective equipment
VTM: viral transport media
LoD: limit of detection
EUA: Emergency Use Authorization
Ct: cycle threshold
GCE: genomic copy equivalents

