## Supplemental Info, Table 1, and Fig 1 for "Polyester Nasal Swabs Collected in a Dry Tube are a Robust and Inexpensive, Minimal Self-Collection Kit for SARS-CoV-2 Testing"

### Supplemental Data

#### Materials and Methods

Swab Physical Specifications and Specimen Absorption Measurements

An electronic digital caliper was used to measure the swab bud width and length for each swab type. To measure width, the caliper was zeroed and then extended to fit the widest portion of the swab bud. The length of the swab bud was measured from the tip of the swab bud to the stem. For each swab type, the average measurement for ten swabs was calculated.

To measure swab absorption, swabs were submerged into a 2 mL tube containing 200 µL PBS for 10 seconds (passive) and then removed or were abraded in a circular motion against the side of the tube five times in each direction and then removed. The tubes were quickly spun down and the absorption volume was calculated from the residual volume in the tube, which was measured with a micropipette.

Quantification of SARS-CoV-2 Specimen Pools

SARS-CoV-2 GCE values were determined for the master positive pool, the low-positive clinical pool (2x LoD), and the high-positive clinical pool (10x LoD) using digital PCR (dPCR) and standard curve methods. For dPCR, three independent RNA extractions from the master positive pool were converted to cDNA using SuperScript IV First-Strand Synthesis System (Fisher Scientific). Following cDNA synthesis, dPCR was performed using the U.S. CDC 2019-nCoV_N2 Assay (Integrated DNA Technologies) and the QuantStudio 3D Digital PCR System to determine the number of SARS-CoV-2 *N* gene copies present in the master positive pool. The low- and high-positive clinical pool viral concentrations were below the dPCR limit of detection, therefore, the *N* gene copies were calculated based on the master positive pool *N* gene values and the dilution factors used to create the low- and high-positive clinical pools. In addition, SARS-CoV-2 viral GCE values were confirmed in the low- and high-positive clinical pools using the standard curve method. Heat inactivated 2019 Novel Coronavirus was obtained from ATCC (#VR-1986HK; lot#70035039: 3.75 x $10^5$ GCE/µL) and analyzed using the TaqPath COVID-19 Combo Kit. A 1:10 serial dilution of the inactivated ATCC viral extraction was used to generate a standard curve: 100,000 to 0.1 GCE/PCR reaction. The low- and high-positive clinical pools were tested at the same time and the GCEs were extrapolated from the standard curve.

**Results**

Taken together, all three swab types (dry polyester, dry foam, and polyester in saline) are stable for up to 48 hours in the absence of cold chain. The SteriPack polyester swabs have also demonstrated comparability to the polyester swabs manufactured by other vendors in all performance evaluations conducted. All of the data from these stability studies has been deposited in a file with the FDA, which allows *in vitro* diagnostics (IVD) manufacturers and laboratories to utilize the 48-hour stability claim for nasal swabs without the requirement to replicate the studies.

**Supplemental Table 1.** Comparison of Copan and SteriPack (#60564) dry polyester swabs using high-positive SARS-CoV-2 specimen pool and human clinical matrix to demonstrate comparability of performance and to validate stability in human matrix. ΔCt: mean Ct stability time point – mean Ct 0 hour.

| Target | Paired Swab | Refrigerated | | | Elevated Temperature | | |
|---|---|---|---|---|---|---|---|
| | | 0 h Mean Ct (n=10) | 72 h Mean Ct (n=10) | ΔCt | 0 h Mean Ct (n=3) | 48 h Mean Ct (n=10) | ΔCt |
| *N* | Copan | 28.4 | 29.3 | 0.9 | 29.5 | 28.7 | -0.8 |
| | SteriPack | 28.3 | 29.2 | 0.9 | 29.6 | 28.4 | -1.2 |
| *ORF1ab* | Copan | 27.6 | 28.1 | 0.5 | 26.5 | 28.4 | 1.9 |
| | SteriPack | 27.4 | 28.1 | 0.7 | 26.6 | 27.9 | 1.3 |
| *S* | Copan | 28.6 | 28.9 | 0.3 | 27.2 | 27.4 | 0.2 |
| | SteriPack | 28.4 | 28.8 | 0.4 | 27.2 | 27.1 | -0.1 |
| *RNase P* | Copan | 25.4 | 26.2 | 0.8 | 23.7 | 25.9 | 2.2 |
| | SteriPack | 25.7 | 26.5 | 0.8 | 24.5 | 26.0 | 1.5 |

**Supplemental Figure 1.** Polyester (poly) and foam nasal swab specimen stability study design with refrigeration (**A,** control arm) and with extended periods of time at high temperatures (**B**, experimental arm). Swabs spiked with the pool of negatives (n=2 per swab type) are not shown.

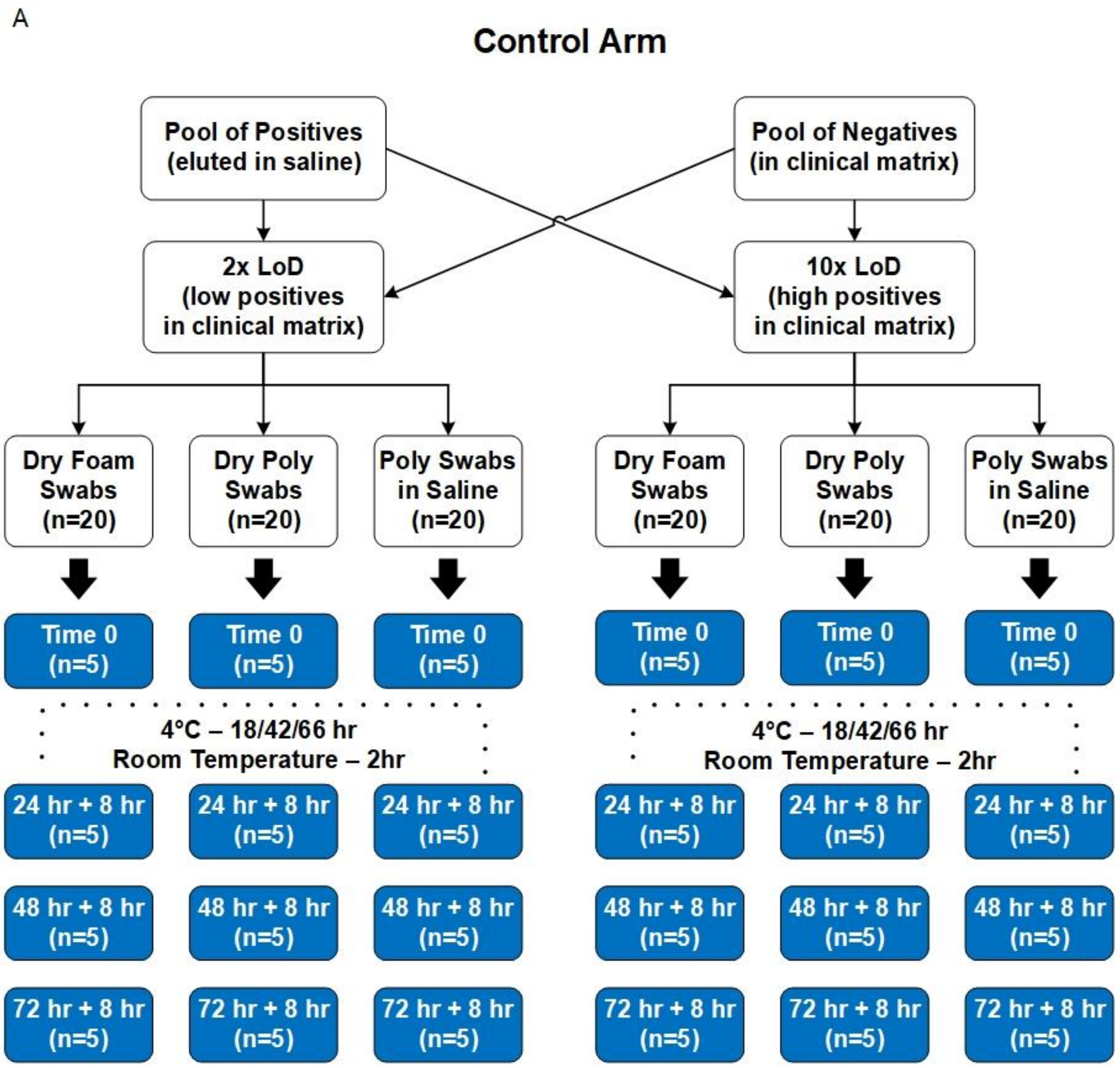

B

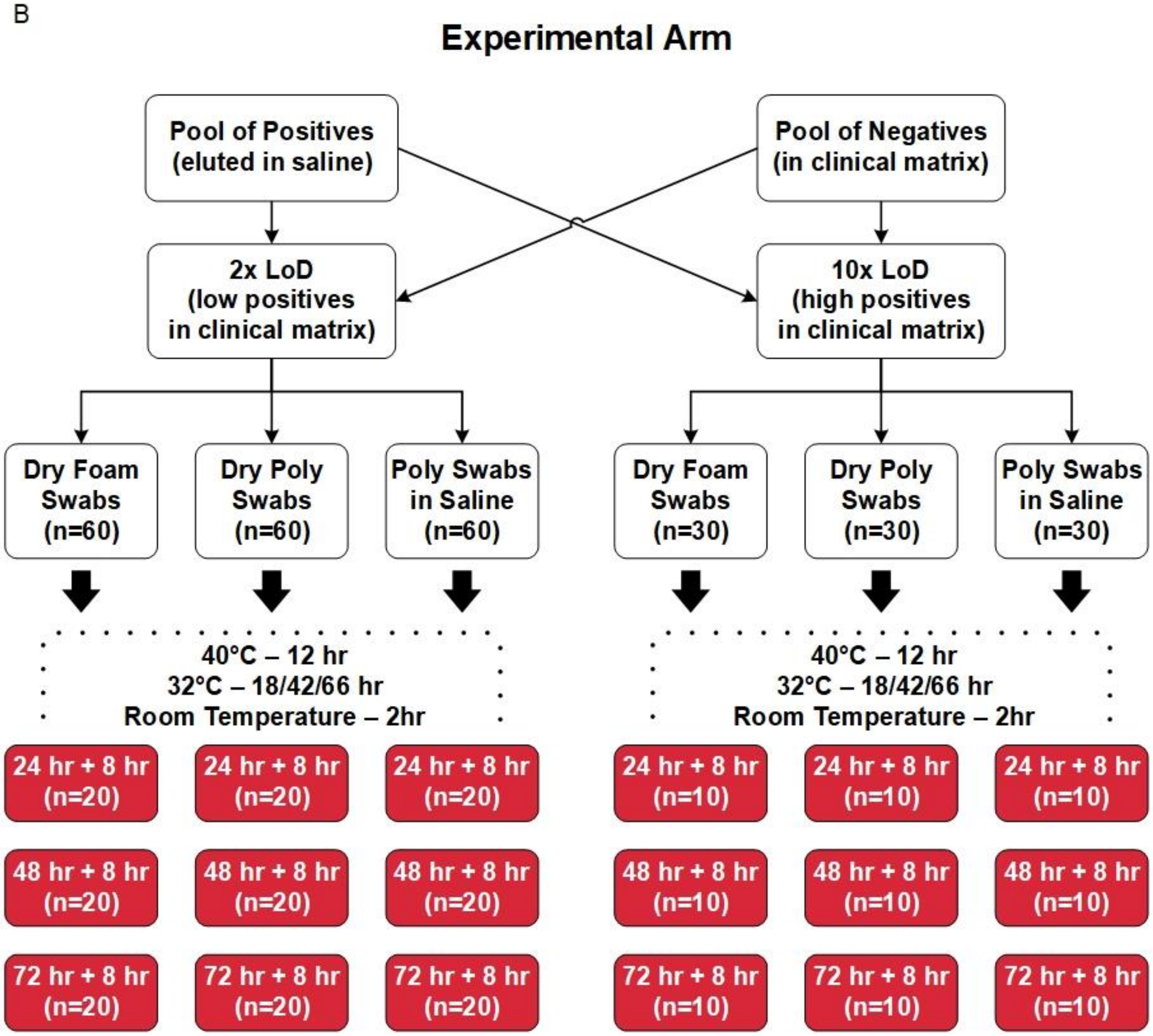
Experimental Arm
Pool of Positives (eluted in saline)
Pool of Negatives (in clinical matrix)
2x LoD (low positives in clinical matrix)
10x LoD (high positives in clinical matrix)
Dry Foam Swabs (n=60)
Dry Poly Swabs (n=60)
Poly Swabs in Saline (n=60)
Dry Foam Swabs (n=30)
Dry Poly Swabs (n=30)
Poly Swabs in Saline (n=30)
40°C – 12 hr
32°C – 18/42/66 hr
Room Temperature – 2hr
40°C – 12 hr
32°C – 18/42/66 hr
Room Temperature – 2hr
24 hr + 8 hr (n=20)
24 hr + 8 hr (n=20)
24 hr + 8 hr (n=20)
24 hr + 8 hr (n=10)
24 hr + 8 hr (n=10)
24 hr + 8 hr (n=10)
48 hr + 8 hr (n=20)
48 hr + 8 hr (n=20)
48 hr + 8 hr (n=20)
48 hr + 8 hr (n=10)
48 hr + 8 hr (n=10)
48 hr + 8 hr (n=10)
72 hr + 8 hr (n=20)
72 hr + 8 hr (n=20)
72 hr + 8 hr (n=20)
72 hr + 8 hr (n=10)
72 hr + 8 hr (n=10)
72 hr + 8 hr (n=10)
